# Genetic support of a causal relationship between Iron status and atrial fibrillation: a Mendelian randomization study

**DOI:** 10.1101/2021.11.02.21265752

**Authors:** Tianyi Wang, Jun Cheng, Yanggan Wang

## Abstract

**Background:** Atrial fibrillation is the most common arrhythmia disease.Animal and observational studies have found a link between iron status and atrial fibrillation. However, the causal relationship between iron status and the risk of atrial fibrillation may be biased by confounding and reverse causality.The purpose of this investigation was to use Mendelian randomization (MR) analysis, which has been widely appied to estimate the causal effect,to reveal whether systemic iron status was causally related to atrial fibrillation.

**Methods:** Single nucleotide polymorphisms (SNPs) strongly associated (P < 5×10^−8^) with four biomarkers of systemic iron status were obtained from a genome-wide association study involving 48,972 subjects conducted by the Genetics of Iron Status consortium.

Summary-level data for the genetic associations with atrial fibrillation were acquired from AFGen (Atrial Fibrillation Genetics) consortium study(including 65,446 atrial fibrillation cases and 522,744 controls). We used a two-sample MR analysis to obtain a causal estimate, and further verified credibility through sensitivity analysis.

**Results:** Genetically instrumented serum iron [OR:1.09;95%; confidence interval (CI)1.02-1.16; *p*=0.01], ferritin [OR:1.16;95%CI:1.02-1.33; *p*=0.02], and transferrin saturation [OR:1.05;95%CI:1.01-1.11; *p*=0.01] had positive effects on atrial fibrillation. Genetically instrumented transferrin levels [OR:0.90;95%CI:0.86-0.97; *p*=0.006] was an inverse correlation with atrial fibrillation.

**Conclusion:** In conclusion,our results strongly elucidated a causal link between genetically determined higher iron status and increased the risk of atrial fibrillation.This provided new ideas for clinical prevention and treatment of atrial fibrillation.

## Backgroud

Atrial fibrillation is the most common arrhythmia in clinical practice^1^.It is associated with 5-fold the risk of stroke and accounts for 15% of all stroke causes^2^.Besides,it is associated with 2-fold the risk of all-cause mortality^3^,affecting the patient’s quality of life and increasing their economic burden. Iron is one of the essential elements for the human body^4^.It plays an important role in physiological processes, such as in oxygen transport, immune function, electron transfer, energy production, DNA synthesis and so on^4^. Furthermore,iron could catalyzes the formation of reactive oxygen species(ROS) and inflammatory factors, which may affect the occurrence of atrial fibrillation^5^.

Some researchers have discovered that even if the left ventricle function properly, atrial fibrillation can occur regardless of the iron status^6^.On the other hand, there are some evidences that iron overload significantly increase the chance of atrial fibrillation^7, 8^.However, inference of observational studies is limited by residual confounding, reverse causation, and detection bias^9^. Therefore, the link between iron status and the risk of atrial fibrillation still needs more attention.

Mendelian randomization uses genetics as an instrumental variable for exposure. While overcoming the limitations of traditional epidemiological researchs, it also strengthens causal inferences about the impact of specific exposure factors on the results^10^. Alleles are randomly distributed during gametogenesis, and genetic variations are randomly distributed in fertilized eggs. The genetic variation precedes the lifestyle and environmental factors selected by the individual, which minimizing the interference of confounding factors, and overcoming the influence of reverse causality^11^. There are no MR-based studies to detect the relationship between iron status and the risk of atrial fibrillation. Therefore, we use the public data of genome-wide association studies (GWASs) to investigate whether iron status may be causally related to the risk of AF.

## Methods

### Study Design Overview

We conducted a two-sample MR study to investigate whether iron status acts as risk factors or mediators in the relationship with atrial fibrillation. Summary data comes from GWAS consortia studies. The original studies were conducted with the informed consent of the participants, as well as ethical approval. We used SNPs as instrumental variables for the iron status. Key assumption methods include the following:(1) SNPs are related to the iron status (the exposure), (2) SNPs are independent of confounding factors, (3) SNPs impact atrial fibrillation (the outcome) only through the iron status(the exposure)^12^.The overall study design is depicted graphically in Figure1.

**Figure.1.**
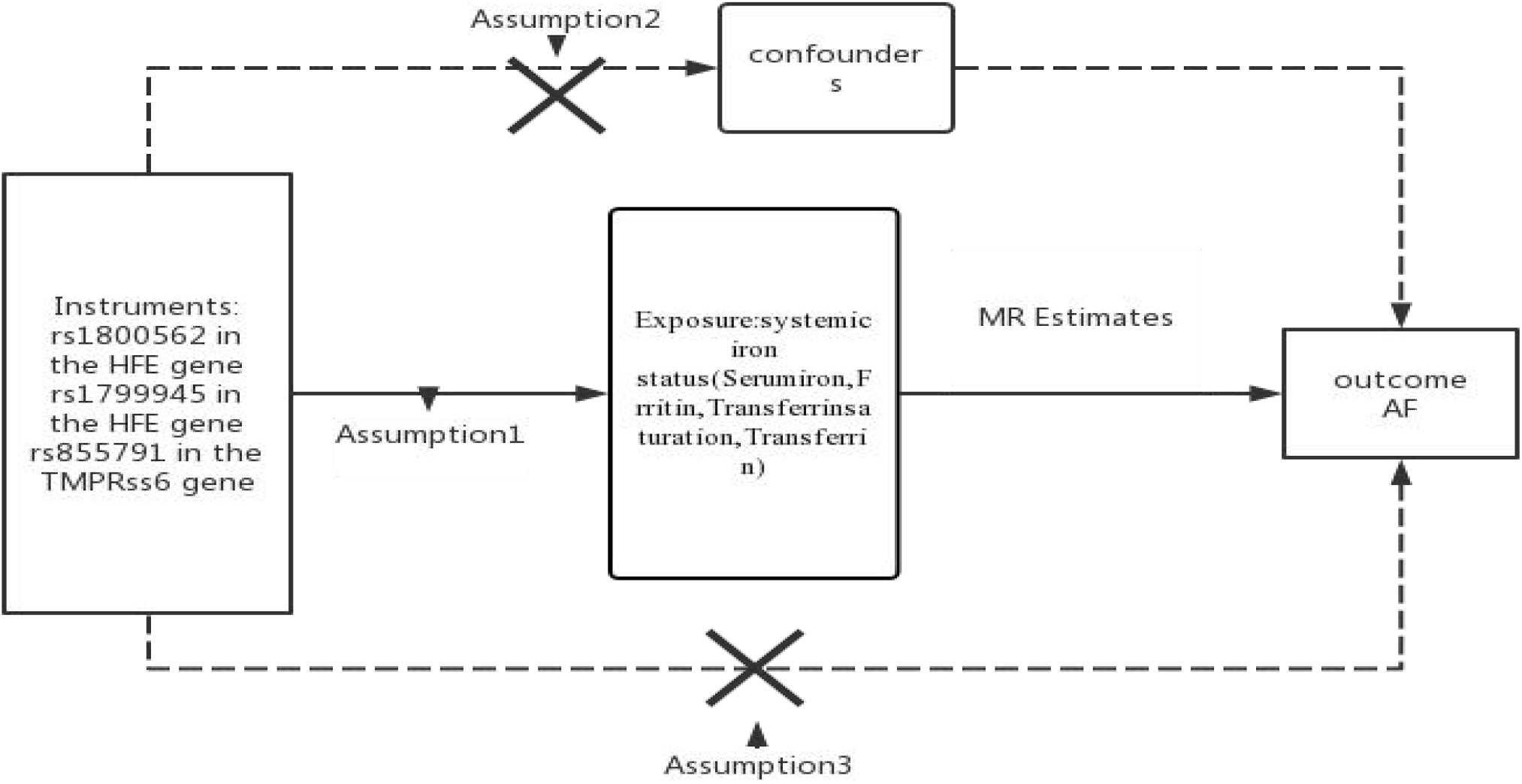
Flowchart of the instrument variables assumptions for MR design. Assumption 1: Genetic variation (SNPs) are closely related to iron status. Assumption 2: Genetic variation (SNPs) are independent of the confounding factors. Assumption 3: Genetic variation (SNPs) affects atrial fibrillation only through iron status

In order to ensure the validity of the instrumental variables for MR analysis, the selection of instrumental variables needs to observe the following standards: Single nucleotide polymorphisms (SNPs) are tightly related to the exposure of the genome-wide significance threshold(p < 5 × 10^−8^)^13^;linkage equilibrium is another necessary requirement for all the SNPS(pairwise r2≤ 0.01)^14^. Weak instrumental variables can also be associated with exposure factors, which can produce an impact on MR research and lead to biases. The generation of weak instrumental variables are generally caused by insufficient sample size. In general, some scholars proposed to evaluate the effect of weak instrumental variables through *F* statistical variable. From the perspective of traditional experience, it is generally better to have an *F*-statistic greater than 10, and it is certainly better to have an *F*-statistic greater than 100. When *F* statistic variables is less than 10, we usually consider that using genetic variation is weak instrumental variable, which may produce certain bias. Hence,we need very careful to interpret the results. Although some scholars believe that *F* statistic may not be a very good instrument to evaluate the bias of weak instrumental variables, we still use *F* statistic at this stage, after all, it is widely used and proved to be a good method, while other new methods still need to be tested in practice. The formula for calculating *F* statistic is as follows: *R*^*2*^*× (N − k − 1)/ [(1 – R*^*2*^*) ×k]*. Here *N* for exposing GWAS research sample, *k* represents the number of independent variable (IV), *R*^*2*^ is IV explain exposed degree coefficient of regression equation (decision). In a two-sample Mendelian randomization study, it is easy to get the specific values of *N* and *K*, but *R*^*2*^ is not easy to get, and we often need to refer to the original literature or the complete GWAS Summary file to get it^15^.

### Genetic associations with systemic iron status

To get the summary-level data on the association between SNPs and iron status, the Genetics of Iron Status (GIS) consortium conducted a meta-analysis of the largest genome-wide association studies (GWASs), including 11 discovery cohorts and 8 replication cohorts and a total of 48,972 European desent (46.9% for male participants) were involved in the meta-analysis. This meta-analysis included a total of 19 cohorts,identified 12 SNPs related to the biomarkers of systemic iron status at genome-wide significance (p<5×10^−8^)(Table 1) and no linkage disequilibrium (LD) among them (all pairwise r^2^≤ 0.01).Five single nucleotide polymorphisms (SNPs) associated with serum iron and transferrin saturation, six SNPs associated with ferritin and eight SNPs associated with transferrin^16^.The increasing of systemic iron status loading means concentration of the transferrin saturation, serum iron and ferritin increasing, while the transferrin decreased^17^.Three of these 12 SNPs,rs1800562 and rs1799945 in HFE and rs855791 in TMPRSS6,showed a concordant change of four biomarkers of systemic iron status at genome-wide significance and accounted for most of the differences in each iron status biomarker^18^. Therefore, these three SNPs have sufficient effects to act as instrumental variables of iron status^19-21^.It seems unlikely that there are biases due to the effect of the weak instrumental variables, because the value calculated by the *F* statistic variable is from 39 to 3,340^22^.The relationship between these SNPs and iron status biomarkers was obtained after the adjustment of covariables, including age and principal component scores, and other study-specific covariates. The details information of SNPS and iron status are presented in in Supplemental Table 1and2.

### Data sources: outcome

Summary-level data through the largest meta-analysis genome-wide association studies for AF(atrial flutter, paroxysmal AF, and persistent AF grouped together) were obtained from the AFGen (Atrial Fibrillation Genetics) consortium study conducted by Roselli C et al in 2018^23^.To ensure that the effect assessments were consistent with same alleles, they using the default settings for the harmonize data command in a two sample MR package in R. This study contained information from more than 50 studies (84.2% European, 12.5% Japanese, 2% African American, and 1.3% Brazilian and Hispanic), including participants from UK Biobank, Biobank Japan and included 65,446 atrial fibrillation cases and 522,744 controls.. Through these two websites (http://afgen.org) and (http://www.kp4cd.org/datasets/v2f) can obtain the data summary statistics and download. The three instrumental varibles SNPs related to iron status were available in the atrial fibrillation outcome GWAS.

### MR Estimates

We mainly used the two-sample MR method to infer the causal association between iron status and AF. Therefore, we needed to use “Two sample MR” in R package (version 0.4.23) to conduct MR analysis. Specifically, the inverse-variance-weighted (IVW)^24^,MR-Egger regression^25^, weighted median, simple mode and weighted mode methods^26^,were used to estimate the effect value between iron status and AF respectively.Weighted median was the primary method to assess the association of genetically predicted iron status and AF risk. In addition, we present the results of different statistical methods in the same chart. The data we need for exposure and outcome were publicly available and accessible in the GWAS database. By using different methods, we could finally get the three main results of Beta, *SE* and *P* value through MR analysis. According to these results, we can calculate 95% confidence interval (CI) and odds ratios(OR) value respectively. The formula required was as follows: OR = exp (beta); CI = exp (beta ± 1.96× SE). During the analysis, there were exposures and other potential confounders that make evaluations difficult. We can obtain limited information through GWAS catalogue database (https://www.ebi.ac.uk/gwas).Through the retrieval of this website and literature reports, it was found that rs1799945 (HFE gene) has a certain relationship with high systolic and diastolic blood pressure^27^.The link between hypertension and atrial fibrillation is likely to lead to atrial fibrillation by dilating the left atrial diameter^28^. But then, we used the leave-one-out method to test the effect of rs1799945 (HFE gene) on the results.

### Sensitivity Analyses

The purposes of our sensitivity analysis were to identify any potential possible pleiotropic and heterogeneous problems. Because one of the assumptions in the MR analysis was that the instrumental variables could only influence the result(AF) by selecting the exposures(iron status), and not by other confounders^29^. In order to exclude the heterogeneity of instrumental variables such as different experiments, platforms and populations, IVW and MR-Egger methods were used to test the heterogeneity. The value of each SNP and Cochran’s Q statistics under each iron status were displayed through forest plots^30^. In addition, leave-one-out analysis were performed to identify SNPS with larger or non-proportional effects by removing one SNP at a time and recalculating estimates for the entire pool of instrumental variables. We used MR-Egger statistical sensitivity analysis to limit the pleiotropic effect of instrumental variables and make MR analysis more reliable. In the MR-Egger regression, the intercept, as an indicator of the mean multivariate bias, can be freely estimated^31^. Power calculations were based on a method designed for a binary outcome^32^. All of the above analysis were conducted by VERSION 4.1.0 of R.

## Results

### 1. Causal effects of iron status on the risk of AF

In the MR analysis results, which were shown as odds ratios (ORs) for AF per standard deviation (SD) increase in each biomarker of iron status and risk of AF. We found that serum iron [OR:1.09;95%confidence interval (CI)1.02-1.16; *p*=0.01], ferritin [OR:1.16;95%CI:1.02-1.33; *p*=0.02], and transferrin saturation [OR:1.05;95%CI:1.01-1.11; *p*=0.01] had a significant positive effects on AF. However, higher transferrin levels [OR:0.90;95%CI:0.86-0.97; *p*=0.006] was associated with a lower probability of AF, which is indicative of decreased iron status. The relationship between each biomarker of iron status and the risk of AF were shown graphically in Figure 2.

**Figure 2:**
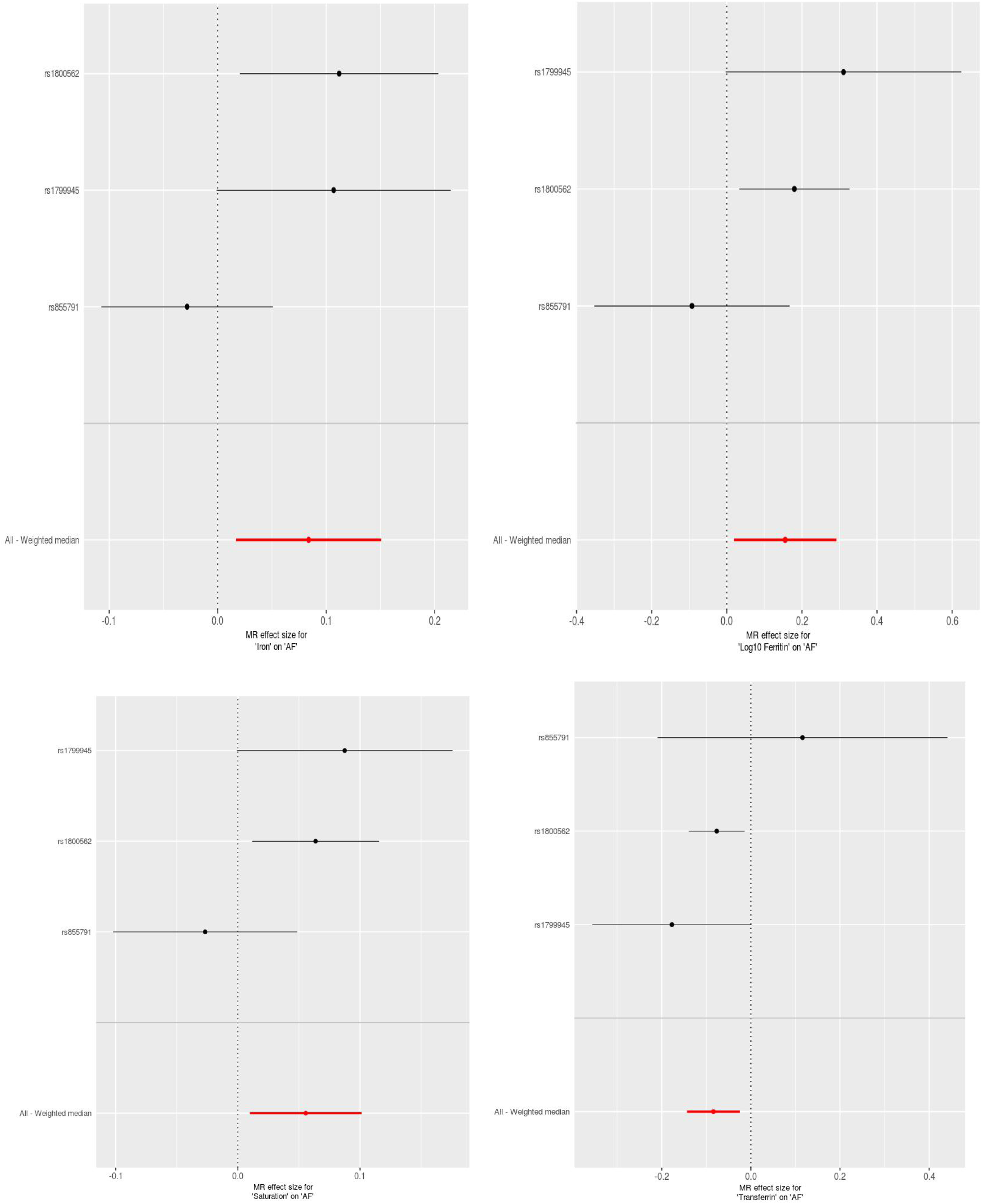
Estimates for the effect of Iron status on AF from two-sample Mendelian randomization. (a) Iron-AF. (b)Log10 Ferritin-AF. (c)Saturation-AF. (d)Transferrin-AF.

### 2. Sensitivity analyses did not display indication of unknown pleiotropy

The presupposition of Mendelian randomization study for causality inference is that there is no level of pleiotropy biases. We used PhenoScanner database to examine the biological pleiotropy of these instruments to evaluate the possible biases^33^. As expected, three SNPS affect Red blood cell traits by changing iron status^34^. The MR-Egger intercepts for the four biomarkers of iron status for directional horizonal pleiotropy did not differ significantly from null (*p*= 0.51, 0.65, 0.55, and 0.79 for serum iron, ferritin, transferrin saturation, and transferrin, respectively). The MR-Egger for the four biomarkers for Heterogeneity test did not differ significantly from null (*p*=0.07,0.07,0.09,0.13 and for serum iron, ferritin, transferrin saturation, and transferrin, respectively) neither. Some articles pointed out that the iron status raising allele at rs1800562 (HFE gene) and at rs1799945 (HFE gene) were association with lower low-density lipoprotein levels and higher systolic and diastolic blood pressures^35^. By searching a large number of existing literatures, there were no clear relationship between hypolipidemia and atrial fibrillation. Although one of the alleles was associated with high blood pressure, which can induce AF,we conducted leave-one-out sensitivity analysis test and found that there were no changes about the MR estimate. Even if the direction of estimates varied somewhat (P > 0.05), but did not change the pattern of results in Figure 3.

**Figure 3.**
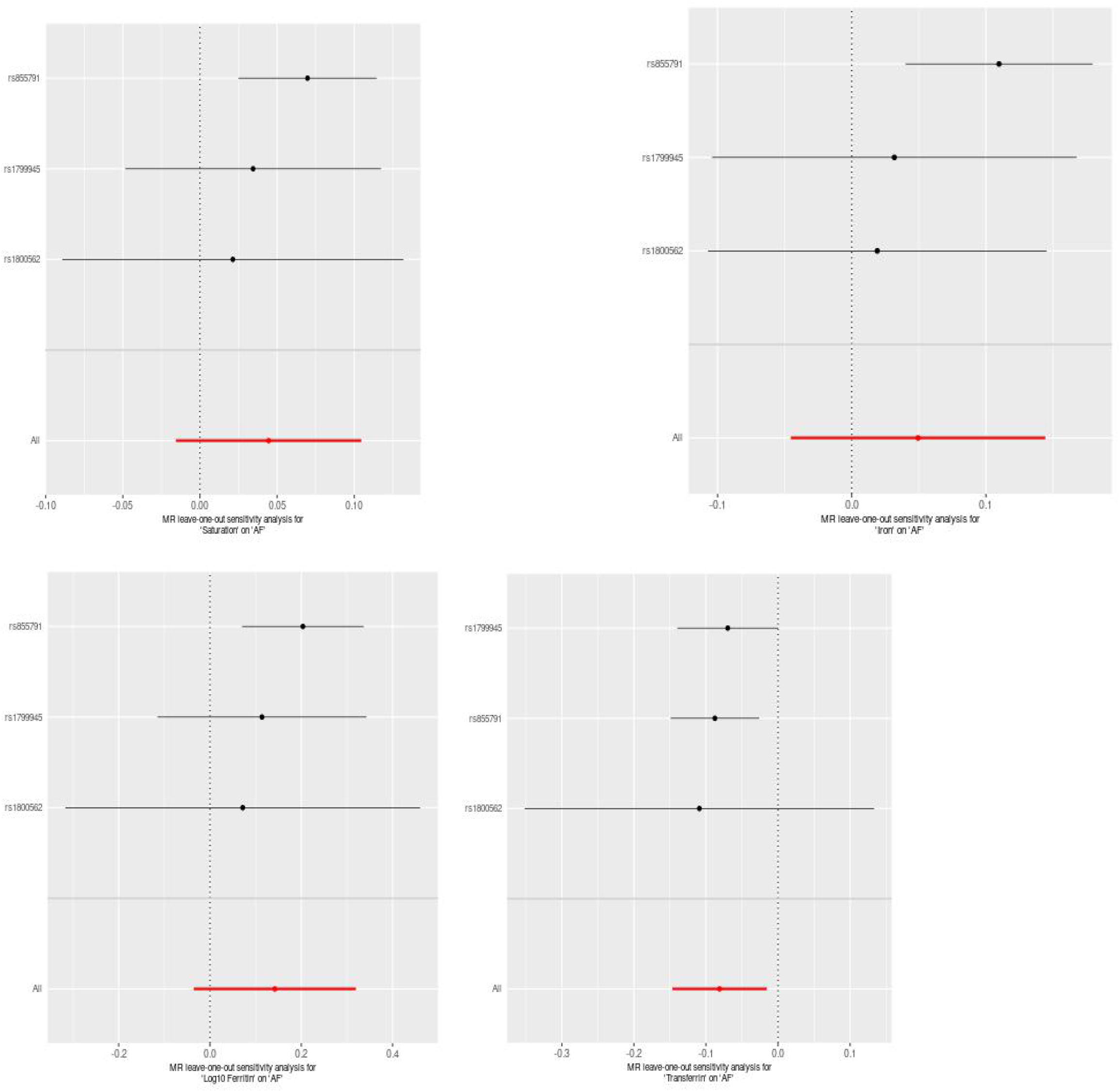
Plots of “leave-one-out” analyses for MR analyses of the causal effect of Iron status on AF in replicative practice. A. Iron-AF. B Log10 Ferritin-AF. C. Saturation-AF. D.Transferrin-AF

MR analysis were carried out by five methods.MR Egger, Weighted median, Simple mode, Weighted mode produced directionally consistent effects as the IVW estimates. We compared the five MR analyses in different iron status and charted them in Figure 4.

**Figure 4.**
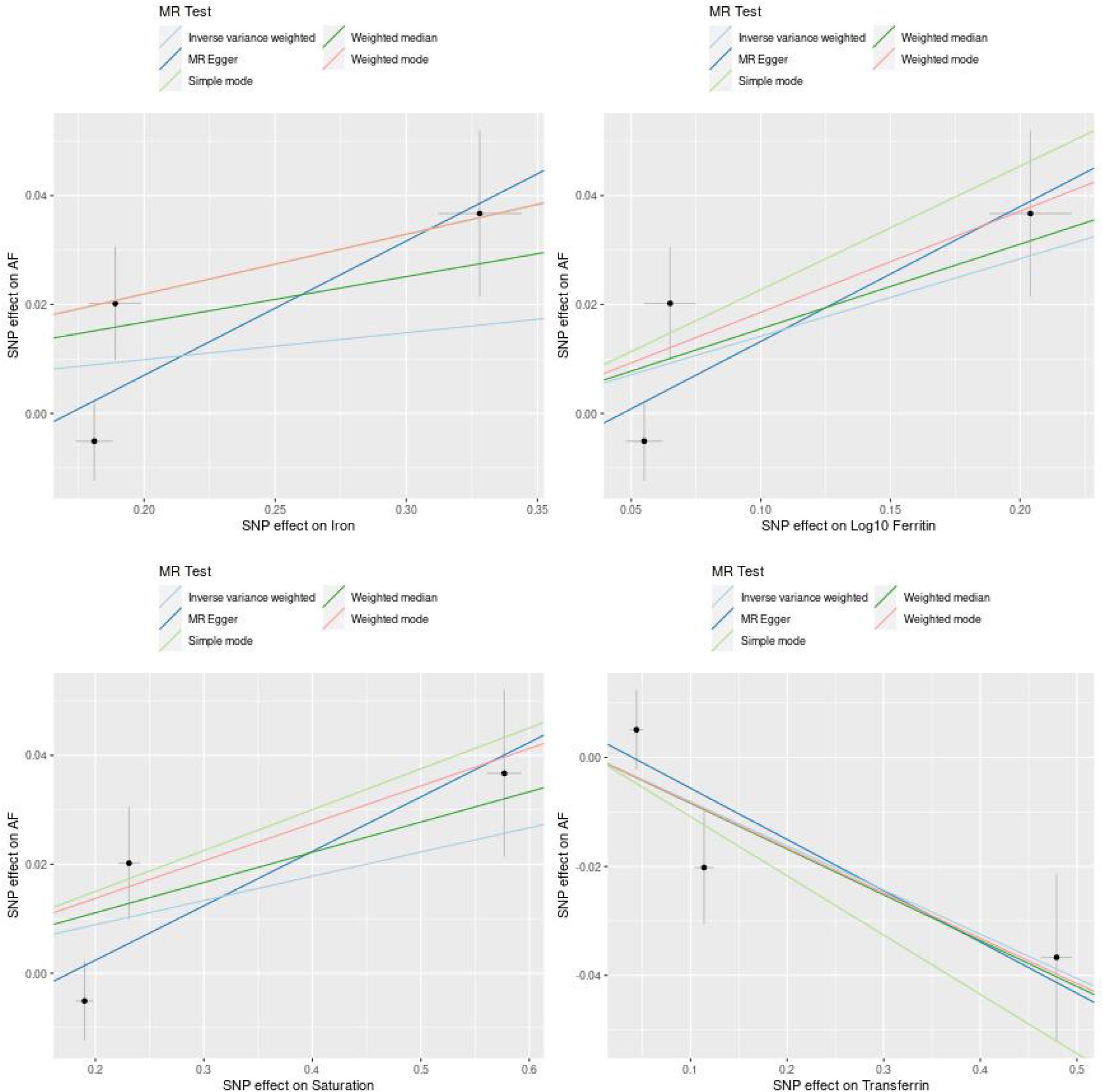
Scatter plots for MR analyses of the causal effect of iron status on AF in initial practice. A Iron-AF. B Log10 Ferritin-AF. C Saturation-AF. D Transferrin-AF. The slope of each line corresponding to the estimated MR effect per method.

## Discussion

Before the time, there have been observational studies of iron status in relation to arrhythmias and atrial fibrillation^36^.Excessive or lack of nutrients can lead to many chronic diseases^37^.Due to the differences in race, ethnicity, sample size, as well as some unconsidered confounding factors or unknown risk factors, the observation results are easy to exist biases. Consequently, we use two-sample MR, which can minimize the interference of confounders, to estimate the associations between several genetically determined markers of systemic iron status(serum iron, ferritin, transferrin saturation, and transferrin) and the risk of AF. The basic assumption about two-sample MR is that the instruments (SNPs) should be associated with the outcome (AF) only via the exposure (systemic iron status as reflected by the four iron biomarkers).Furthermore, we use summary level data generally mainly from the largest meta-GWASs of European descent to reduce the likelihood of biases^38^.The final results showed that iron overload is associated with an increased risk of AF based on MR analysis. Moreover, the conclusions obtained by different MR analysis methods are consistent. These results provide a reasonable way for using iron status as a promising clinic target for AF prevention and treatment^39^.

The reliability of the results may be not stable due to the pleiotropic effects of MR analysis method^40^.We used SNPS in PubMed database to search for the possibility of the secondary phenotypes. The association of the 3 iron status instruments are well-established relationship between iron status and anemia^16^. But if any effect of RBC traits on AF risk was downstream of iron status, instead of independent of it, this would not bias the MR analysis^41^.Our online search identified two SNPS —rs1800562 and rs1799945 in HFE gene. The former is associated with lower low-density lipoprotein levels, the latter is associated with higher systolic and diastolic blood pressures. The influence of blood lipids on atrial fibrillation have not been clearly reported. However, blood pressures have been reported to increase the risk of AF^28^. By causing left ventricular hypertrophy, atrial dilatation is more likely to lead to atrial fibrillation^42-44^.

Nevertheless, removing the SNP produced no substantive effect in MR analysis results, suggesting that the MR estimates in the present study were not likely expected to be biased by blood pressure. Due to the influence of different MR analysis methods, the slight differences in the confidence interval width and estimates may be explained by accidental, possibly, or results of a difference measurement errors, rather than indicating distinct real differences. In addition, using the MR-Egger method to conduct the pleiotropic test did not detect biases and associationes. Both exposure and outcome in the public genome wide association study database mainly from European population, thus minimizing the biases of population stratification. Meanwhile, the leave-one-out MR estimate was parallel to the primary MR estimates. Generally speaking, it shows that there were unlikely serious biases in our research methods and conclusions.

Iron deficiency and systemic iron overload can cause metabolic disorders. About one-third of patients with heart failure and one-half of patients with pulmonary hypertension have iron deficiency. Moreover, iron deficiency has an adverse effect on patients with coronary artery disease, heart failure, pulmonary hypertension, and patients who may undergo heart surgery^45^.Systemic iron status increasing means that serum iron, transferrin saturation and ferritin levels rise, while transferrin levels decreases^46^.Previous MR studies have shown that increased iron status reduces the risk of coronary artery related diseases^47^and Parkinson’s syndrome^48^ and increased the risk of type 2 diabetes^49^ and cardiogenic thrombosis^21^.As far as we known, no causal relationship have been conducted previously between instruments iron status and atrial fibrillation using the MR methods. Hence, we suggested a casual relation between systemic iron status and AF for the first time.

The most important risk factors for the occurrence of atrial fibrillation events are aging, hypertension diabetes, atrial dilation and left atrial enlargement, stroke, cardiomyopathies, heart failure and genetics^50^.In addition, natriuretic peptide levels and volume overload also increase the probability of atrial dilation and atrial fibrillation^51^.Our MR study found that one of the alleles related to iron status at rs1799945(HFE gene) which is related to high systolic blood pressure and high diastolic blood pressures^27, 52^.As mentioned above, hypertension is one of the risk factors for atrial fibrillation, which is as a kind of confounding factor leading to the effect of iron status on AF. Through leave-one-out pleiotropic analysis, the results did not change significantly. When iron overload is determined by the cardiac magnetic resonance (CMR) value, the risk of atrial fibrillation is significantly increased^53^.Whether there are potential mechanisms that make the iron overload directly affect the occurrence of AF. We could get clues from some observations and basic experimental results.

First of all, in addition to the above-mentioned volume overload is the main reason for atrial dilation, another major factor is oxidative stress^54^.For all we know, iron overload is an important cause of oxidative stress^55^.Oxidative stress can induce changes in intracellular calcium ions, lead to delay depolarization, result in atrial focal ectopic action potential^56^.In addition, abnormal calcium ion handling and calcium overload could induce cardiac remodeling, resulting in atrial fibrillation^57^. When the iron overload exceeding the iron storage capacity, erratic iron enters the circulation^58^ and can also penetrate into the myocardial cells^55^.Iron overload cardiomyocytes have abnormal action potentials comparing with normal cells. Animal experiments have found that iron toxicity can lead to change in electrical conduction of the heart and arrhythmia^59^.Iron toxicity damages cardiomyocytes conduction still by increasing oxidative stress. Oxidative stress breaks the balance of sodium, potassium, and calcium channels, resulting in abnormal cell ion flow, insufficient atrial contraction and atrial fibrillation^55.^Another mechanism is that oxidative stress activates Nuclear factor-κB (NF-κB). NF-κB can down-regulates calcium channels and lead to atrial fibrillation^60^.There is an experiment on older rats, the result is that iron overload can cause oxidative stress and activate NF-κB in brain tissue^58^.Besides,excess free ironions can participate in the Fenton reaction and produce a large amount of OH^-^,which is considered to be the strong activtion of ROS^61, 62^. In 2012, Dixon et al discovered a new way of programmed cell death—ferroptosis, the essence of this is the Fenton reaction^63^.Thus, it can be fully inferred that iron overload may cause atrial fibrillation through ferroptosis.It had been proved in animal experiments that ferroptosis occurs in atrial fibrillation, and inhibiting ferroptosis can effectively control the occurrence of atrial fibrillation^64^. Finally, some studies have found that in thalassemia patients, iron overload is caused by ineffective hematopoiesis and repeated blood transfusions^65, 66^.In these patients, inflammatory factors are elevated. By using iron chelating agents, the level of inflammatory factors demonstrate a downward trend^67^. So the iron overload can cause changes in the heart through inflammatory factors, making the heart more prone to atrial fibrillation, and it is one of the regulators of inflammation^57^. In summary, iron status can lead to atrial fibrillation through potential mechanisms: oxidative stress, inflammatory response, and ferroptosis. Our findings provided the first evidence that four genetically determined systemic iron status biomarkers (serum iron, ferritin, ferritin saturation, ferritin) were significantly associated with atrial fibrillation risk.That was consistent with some previous recognized association between iron and AF based on observational studies.

Herein, our researches should be interpreted in conbination of some limitations. First, due to our analyses were conducted at publicly available GWAS databases, they were difficult to perform stratified analysis such as age and gender in the exposures and out come databases. Furthermore,even though we used different methods to try to minimize the pleiotropy and data obtaining from the GWAS database with the largest sample size in the world, biases caused by unknown biological effects of the SNP on iron status may be inevitable. Moreover, the datasets used in our research were mainly derived from European races, which aiming at reducing the bias owing to races. Hence, it is unclear whether the results certainly appropriate for other races. Finally,this MR study investigated the relationship between iron states and in the normal range of AF.For this reason,therefore, our results cannot be used to infer the effects of abnormally high serum iron levels caused by veins, long-term oral iron supplementation, or hemochrome disease.

Ultimately, some Mendelian Randomization studies used IV-W as the main method of analysis, while we used weighted median for the following reasons, in order to make the results more reliable and reasonable.As a valid IV, there are three necessary assumptions: IV1: the variant is predictive of the exposure;IV2: the variant is independent of any confounding factors of the exposure-outcome association;IV3: the variant is conditionally independent of the outcome given the exposure and the confounding factors^68^. Of these, only IV1 can be verified(p<5×10^−8^), while the rest depends on all possible confounding factors of exposure and outcome,both measured and unmeasured.As Mendelian randomization analysis contains multiple variants, statistical capacity is improved^69^. But one of the challenges is that not all included genetic variants are valid IVs^70^.If all genetic variants satisfy the IV assumptions, IVW can adequately reflect the real causality. However, when an invalid IV occurs, the IVW deviates from the true causality, resulting in a bias outome^71^. It’s like that Voight et al suggested there is not even a moderate causal effect of HDL-c on CAD risk^72^. In this study, we found that one of the IV may be an invalid or weak instrument (it is associated with the confounding factor of hypertension),so using the IVW method is not appropriate. The weighted median methods generally have more power with a positive causal effect, especially when the proportion of invalid IVs increases and also have less mean standard errors than the IVW method^71^.Therefore, for this study, weighted median methods are more statistically close to the real causality.

## Conclusion

Through the MR method, we tested the previously assumed hypothesis that there is a causal relationship between the overload in the iron status of the systematic and the occurrence of AF. We relied on iron status data measured in 48,972 individuals in the general population database and the AF Gen (Atrial Fibrillation Genetics) consortium study including participants from UK Biobank, Biobank Japan and included 65,446 atrial fibrillation cases and 522,744 controls in the general population to conduct a two-sample MR experiment. We used the three SNPs as instruments to increase statistical power by combining their MR estimates and investigate possible pleiotropic effects. Our Mendelian randomization study first showed that systemic iron status increases AF risk. This has important clinical significance for the treatment of borderline anemia and the continued iron therapy of anemia patients after the iron status is corrected, especially for patients with anemia or continuous supplementation for patients at high risk of atrial fibrillation, careful consideration may be needed.

## Supporting information

Supplemental Table1,Table2,and will be used for the link to the file on the preprint site

## Data Availability

Single nucleotide polymorphisms (SNPs) strongly associated with four biomarkers of systemic iron status were obtained from a genome-wide association study involving 48,972 subjects conducted by the Genetics of Iron Status consortium. Summary-level data for the genetic associations with atrial fibrillation were acquired from AFGen (Atrial Fibrillation Genetics) consortium study(including 65,446 atrial fibrillation cases and 522,744 controls) .

## supplementary information

The following are available online link.Table S1: iron status on genome-wide significance level biomarkers related SNPs, and included in all three main SNPs and secondary analysis of the main analysis;Table S2:Additional information was amplified for three major SNP.

## Author Contributions

T.Y.W.designed the study and wrote the article. J,C. provided technical support. Y.G.W. revised the article.

## Ethics approval and consent to participate

This MR study only uses publicly available data and did not collect original data. It did not have ethical approval because it only used published or publicly available data, which can be found in the original publications.

## Informed Consent Statement

Patient consents were waived due to the use of published or publicly available data for the study. We collected no original information for this MR study. Informed consent from each participant who participates in each of the studies included in the investigation can be found in the previous publications.

## Availability of data and material

All relevant data supporting the conclusions are included in this published article and its supplementary information files.

## Acknowledgements

We thank all members of our research team for their wholehearted cooperation and their excellent work.We also thank Wuhan University Zhongnan Hospital Internal Medicine for its methodological support.

## Consent for publication

Not applicable.

