## Supplemental Table1,Table2,and will be used for the link to the file on the preprint site for "Genetic support of a causal relationship between Iron status and atrial fibrillation: a Mendelian randomization study"

Table1.iron status on genome-wide significance level biomarkers related SNPs, and included in all three main SNPs and secondary analysis of the main analysis.^[1]^

| SNPS CHR Nearby EA EAF  Gene | | | | | Transferrin g/L  Beta SE p | | | Serum ironµmol/L  Beta SE p | | | Transferrin Saturation, %  Beta SE p | | | | Log10 Ferritin,µg/L    Beta SE p | | | |
| --- | --- | --- | --- | --- | --- | --- | --- | --- | --- | --- | --- | --- | --- | --- | --- | --- | --- | --- |
| rs179994* | 6 | HFE | G | 0.15 | -0.114 | 0.010 | 9.4x10^-30^ | 0.189 | 0.010 | 1.1x10^-81^ | 0.231 | 0.010 | | 5.1x10^-109^ | | 0.065 | 0.010 | 1.7x10^-10^ |
| rs180056* | 6 | HFE | A | 0.07 | -0.479 | 0.016 | 8.9x10^-196^ | 0.328 | 0.016 | 2.9x10^-97^ | 0.577 | 0.016 | | 2.2x10^-270^ | 0.204 | | 0.016 | 1.5x10^-38^ |
| rs855791* | 22 | TMPRSS6 | G | 0.55 | -0.044 | 0.007 | 2.0X10^-9^ | 0.181 | 0.007 | 4.3X10^-139^ | 0.190 | | 0.007 | 6.4X10^-137^ | 0.055 | | 0.007 | 1.4X10^-14^ |
| rs8177240 | 3 | TF | G | 0.35 | 0.380 | 0.007 | 8.4X10^-610^ | 0.066 | 0.007 | 6.6X10^-20^ | 0.100 | 0.008 | | 7.2X10^-38^ |  | |  |  |
| rs7385804 | 7 | TFR2 | A | 0.62 |  |  |  | 0.064 | 0.007 | 1.4X10^-18^ | 0.054 | 0.008 | | 6.1X10^-12^ |  | |  |  |
| rs651007 | 9 | ABO | C | 0.79 |  |  |  |  |  |  |  |  | |  | 0.050 | | 0.009 | 1.3X10^-8^ |
| rs174577 | 11 | FADS2 | A | 0.36 |  |  |  |  |  |  |  |  | |  | 0.062 | | 0.007 | 2.3X10^-17^ |
| rs6486121 | 11 | ARNTL | C | 0.34 | 0.046 | 0.007 | 3.9X10^-10^ |  |  |  |  |  | |  |  | |  |  |
| rs4921915 | 8 | NAT2 | A | 0.76 | 0.079 | 0.09 | 7.1X10^-19^ |  |  |  |  |  | |  |  | |  |  |
| rs9990333 | 3 | TFRC | C | 0.53 | 0.051 | 0.007 | 2.0X10^-13^ |  |  |  |  |  | |  |  | |  |  |
| rs744653 | 2 | AC013439.4 | C | 0.16 |  |  |  |  |  |  |  |  | |  | 0.089 | | 0.010 | 8.4X10^-19^ |
| rs411988 | 17 | TEX14 | G | 0.44 |  |  |  |  |  |  |  |  | |  | 0.044 | | 0.007 | 1.6X10^-10^ |

CHR, chromosome;EA indicates effect allele; EAF, effect allele frequency; SE, standard error; SNP , single nucleotide polymorphism. * Three SNPs were included in the main analyses.

Table2.

Additional information was amplified for three major SNP^[1]^

|  | Instruments-Exposure Associations (n=48 972) | | | | | | | | | | | | | | | |
| --- | --- | --- | --- | --- | --- | --- | --- | --- | --- | --- | --- | --- | --- | --- | --- | --- |
|  | Iron, μmol/L | | | | TransferrinSaturation, % | | | | Log10Ferritin,μg/L | | | | Transferrin, g/L | | | |
| SNPS | R^2^ | F | GX | GXSE | R^2^ | F | GX | GXSE | R^2^ | F | GX | GXSE | R^2^ | F | GX | GXSE |
| rs1800562 | 1.3 | 668 | 0.328 | 0.016 | 4.2 | 2127 | 0.577 | 0.016 | 0.5 | 256 | 0.204 | 0.016 | 2.9 | 1446 | -0.479 | 0.016 |
| rs1799945 | 0.9 | 450 | 0.189 | 0.010 | 1.4 | 676 | 0.231 | 0.010 | 0.1 | 53 | 0.065 | 0.010 | 0.3 | 163 | -0.114 | 0.010 |
| rs855791 | 1.6 | 806 | 0.181 | 0.007 | 1.8 | 889 | 0.190 | 0.008 | 0.1 | 73 | 0.055 | 0.007 | 0.1 | 47 | -0.044 | 0.007 |

F, F statistic; GX, the per-allele effect on SD units of the iron marker; GX SE, standard error of GX; R^2^, percentage of the iron marker variation explained by the SNP; and SNP, single-nucleotide polymorphisms.
